# Patient perceptions towards psychedelics for musculoskeletal pain: A cross-sectional survey

**DOI:** 10.64898/2026.05.29.26354422

**Authors:** Erica J. Li, Basmah Mosharraf, Hiba Ali, Mackenzie Noyes, Pranjali Doshi, Christine Wallace, Rotem Petranker, Anthony Adili, Moin Khan, Jason W. Busse, James MacKillop, Kim Madden

**Affiliations:** Department of Psychology, Neuroscience & Behaviour, McMaster University, Hamilton ON Canada; St. Joseph’s Healthcare Hamilton, Hamilton ON Canada; School of Public Health Sciences, University of Waterloo, Waterloo ON Canada; Department of Life Sciences, McMaster University, Hamilton ON Canada; Department of Surgery, McMaster University, Hamilton ON Canada; Department of Anesthesia, McMaster University, Hamilton ON Canada; Department of Health Research Methods, Evidence & Impact, McMaster University, Hamilton ON Canada; Department of Psychiatry & Behavioural Neurosciences, McMaster University, Hamilton ON Canada

## Abstract

**Background:** Psychedelics are emerging as potential management options for chronic musculoskeletal pain due to preliminary evidence of effectiveness and low addictive potential, but patients’ perceptions remain unknown. This study assessed patient perceptions regarding psilocybin for musculoskeletal pain.

**Methods:** We conducted a cross-sectional survey of adults (≥19) with musculoskeletal pain attending a hospital-based orthopaedic clinic. Participants reported demographics, perceptions of psychedelics for pain management, and willingness to participate in psychedelic research. Multivariable regression explored factors associated with perceived analgesic potential, and willingness to try a full therapeutic dose of psilocybin or a microdose.

**Results:** Among 295 participants, 73% reported moderate-to-severe pain; 75% used analgesics; of these, 41% used opioids (86/209). While 24% reported prior psychedelic use, only 3% had discussed psychedelics with a healthcare provider. Most perceived that psilocybin had moderate-to-high effectiveness for pain (76%). Most respondents endorsed a moderate-to-high willingness to try microdoses (58%) and macrodoses (53%) of psilocybin for pain. Prior non-therapeutic psychedelic use predicted a 1.05-unit increase in perceived analgesic potential on the 10-point scale (*p*=.013). Willingness to try a macrodose of psilocybin was most strongly associated with prior non-therapeutic (*B*=3.16) and therapeutic (*B*=2.42) psychedelic use; in contrast, pain severity had a significant but modest association, with a 0.21-point increase in willingness for every 1-unit increase in pain severity (*p*=.017). Similarly, willingness to try a microdose of psilocybin was predicted by non-therapeutic (*B*=2.82) and therapeutic (*B*=2.48) use, whereas the effects of pain severity (*B*=0.20) and younger age (*B*=−0.30) were significant but small. Most respondents (52%) reported moderate-to-high willingness to participate in a trial of psilocybin for pain relief, and health risks were the primary concern (33%).

**Conclusions:** Study findings suggest a majority hold neutral-to-positive perceptions of psilocybin for pain. Addressing perceived barriers, including health effects and gaps in patient knowledge, should be considered when designing future trials.

## INTRODUCTION

The opioid epidemic in Canada remains a critical public health crisis, marked by high rates of overdose deaths and related harms [1]. From October 2024 to September 2025, Canada recorded 5,724 opioid-related deaths (16 daily on average) [2]. The crisis stems from multiple factors: the rise of potent illicit drugs like fentanyl, socioeconomic inequities, untreated mental health issues, limited addiction services, and the over prescription and misuse of opioids [3,4]. Excessive opioid prescribing has played a significant role in the opioid epidemic [5]. Opioids, valued for their potent analgesic effects, are frequently prescribed after orthopaedic surgeries for moderate to severe pain [6]; orthopaedic surgeons are among the top prescribers of opioids [7]. However, the use of opioids carries risks, including long-term use, physiological side effects, altered cognition, and diversion for non-medical purposes [3]. Given these risks and the limited evidence supporting the long-term effectiveness of opioids, there is increasing interest in non-opioid alternatives for perioperative and chronic pain management [8–10].

Psychedelics have emerged as a potential alternative to traditional pain management, utilizing pathways distinct from conventional analgesics to influence perception, mood, and cognition [9,11]. [12–14]. The mechanisms of these substances vary by class: classic psychedelics like psilocybin, lysergic acid diethylamide (LSD), and *N*,*N*-dimethyltryptamine (DMT) primarily act as serotonin 2A (5-HT_2A_) receptor agonists, whereas atypical psychedelics like ketamine function as N-methyl-D-aspartate (NMDA) receptor antagonists with serotonin-dopamine reuptake inhibition properties, and 3,4-methylenedioxymethamphetamine (MDMA) acts primarily as a monoamine releasing agent and reuptake inhibitor [14]. Classic psychedelics are proposed to leverage 5-HT_2A_ receptor activation to alter maladaptive neural connectivity and promote neuroplasticity, while simultaneously modulating descending pain pathways and inflammatory processes as an alternative to traditional analgesics [13,15,16]. Classic psychedelics offer a distinct safety advantage over traditional analgesics. They carry virtually no addictive potential, pose no risk of physical overdose, and do not result in physical dependence or withdrawal symptoms [9].

However, significant barriers to widespread adoption remain. Most psychedelics are not legal for medical or recreational use in much of the world, with the exception of ketamine which is commonly used medically for procedural sedation and anesthesia as well as the treatment of depression [17,18]. The administration of psychedelic-assisted therapy is highly resource-intensive, requiring specialized settings and extended clinical supervision [19]. Microdosing, the use of low, sub-impairing doses, may require fewer resources, though the evidence remains limited, with few randomized clinical trials completed to date [20].

A systematic review found a recent sharp increase in clinical research involving psilocybin, with more than half of all studies published between 1964–2023 occurring in the final period of 2021–2023 [21]. Most research has focused on mental health condition; however, there have been observational studies and small open-label and controlled trials suggesting a possible use for the treatment of migraines [22], cluster headaches [23,24], fibromyalgia [25], phantom limb pain [26], and chronic neuropathic pain [27]. There are currently no published studies that have investigated the effects of psychedelics on musculoskeletal pain management, however there are several ongoing clinical trials on psilocybin for fibromyalgia [28] and low back pain [29].

Musculoskeletal (MSK) disorders, encompassing conditions such as osteoarthritis, chronic back pain, and soft-tissue disorders, impacted approximately 1.68 billion people globally [30] and incurred an estimated $2.1 trillion in annual global economic costs [31]. In Ontario, Canada, where this study was conducted, 28.5% of the adult population sought clinical care for MSK disorders within the 2013–2014 fiscal year [32]. The primary objective of this study was to examine patient experiences, perceptions, attitudes, and beliefs regarding the use of psychedelics for musculoskeletal pain. Our secondary objectives included identifying key barriers to participation in trials involving psychedelics and exploring associations between participant characteristics, their beliefs regarding the analgesic potential of psychedelics, and willingness to use psychedelics for pain relief.

## METHODS

### Study Design

This was a cross-sectional survey that consisted of a patient questionnaire to determine perceptions, attitudes, beliefs, and opinions around the use of psychedelics for musculoskeletal pain.

### Questionnaire Development

We developed our survey by modifying two previous cross-sectional surveys of patient perceptions of cannabis use for musculoskeletal pain [33,34]. We also incorporated questions from a survey that examined knowledge, perceptions, and usage of psychedelics among fibromyalgia patients [25]. Our final survey consisted of 34 items using a 0–10 Likert scale or multiple selection format. The questionnaire collected information on participant demographics, and experience with psychedelics, both for therapeutic and non-therapeutic purposes. Given the absence of a formal medical regulatory framework for these substances, ’therapeutic’ use was defined as use for the management of specific physical or mental health conditions, regardless of whether that use was self-directed or prescribed/authorized by a healthcare professional. ‘Non-therapeutic’ use referred to use for “social, fun, or personal exploration purposes.” Participants were then asked about their perception of psychedelics for pain, willingness to participate in psychedelics-based trials, perceived barriers to using psychedelics for pain management, and overall perceived effectiveness of these interventions for pain. To evaluate interest in a macrodose (standard hallucinogenic dose) application, participants were asked to rate their willingness to try “psilocybin (magic mushrooms) for your pain”. This was contrasted with a microdosing framework, which was defined to participants as “using a very low dose (microdosing magic mushrooms)”. The last section of the survey asked participants about their current pain and previous pain management.

### Setting and Participants

Participants were recruited from the outpatient orthopaedic fracture clinic at St. Joseph’s Healthcare Hamilton (SJHH), in Ontario, Canada from June to December 2025. This setting included a population referred by a primary care and emergency room physicians for a broad spectrum of musculoskeletal conditions, including bone and joint pain. Participants were included if they were aged 19 years or older. This threshold aligns with the legal age for purchasing regulated adult substances (e.g., alcohol and cannabis) in Ontario, ensuring the sample consists of individuals who have reached full legal autonomy regarding substance access and decision-making. Inclusion criteria were being a clinic patient referred for assessment or treatment for musculoskeletal pain; being able to read and understand English sufficiently to complete the survey; and judged to have no cognitive impairments that would impede their ability to provide informed consent.

### Questionnaire Administration

Researchers screened patients for eligibility and obtained informed consent. Because this survey asked about participation in illegal activities, the survey was completely anonymous, and we did not request a name or signature on the consent form. Participants indicated their consent by selecting “Yes” on a digital form approved by the research ethics board (REB). Eligible participants then completed the anonymous questionnaire via tablet in REDCap (Vanderbilt University, Nashville TN) [35].

Participants were allowed to withdraw at any time before submission of the survey. All procedures were approved by the Hamilton Integrated Research Ethics Board (HiREB #18196), and the study was conducted in accordance with the ethical standards outlined in the Declaration of Helsinki.

### Sample Size

The sample size was calculated using a standard formula for estimated proportions. We assumed an expected proportion of 0.2, based on population data indicating a lifetime psychedelic use rate of 16% in Canada [36]. This value was rounded up to 20% to provide a more conservative estimate, accounting for potential underreporting or higher prevalence rates within the local population. With a 95% confidence interval, and a 5% margin of error (*d*=.05), the calculation yielded a target sample size of 246 patients. Taking potential incomplete surveys into consideration, we increased the sample size by 20%, resulting in a final target sample size of 295 patients.

### Data Analysis

All statistical analyses were completed in R (v4.2.1; R Core Team, 2022) [37]. We generated frequencies for all demographic information (age, gender, education, income, and race). Likert ratings were classified into low (0-3), moderate (4-6), and high (7-10) for attitudes, and mild (0-3), moderate (4-6), and severe (7-10) for pain severity. These bucketed Likert ratings were used for descriptive reporting to facilitate clinical interpretation. Perceived analgesic effectiveness was measured from 0 to 10 on a continuous scale. Means and standard deviations (SDs) were used for continuous data and frequencies and percentages used for categorical data.

An exploratory paired t-test evaluated intra-individual differences between willingness to try macrodose versus microdose psilocybin, and between the perceived psychological versus physical risk of psilocybin. Independent samples t-tests evaluated differences in attitude ratings stratified by prior psychedelic use and, subsequently, by pain severity (low pain vs. moderate-to-high pain).

To evaluate the relationship between predictor variables and patient attitudes, we constructed three separate Ordinary Least Squares (OLS) linear regression models. The value of the unstandardized regression coefficient represents the change in response score on the 0-10 Likert scales for each dependent variable. The dependent variables were: 1) perceived analgesic potential of psychedelics, 2) willingness to try a macrodose of psilocybin for pain, and 3) willingness to try a microdose of psilocybin for pain. Each model adjusted for clinical and demographic covariates. The regression equation was defined as described in (1).

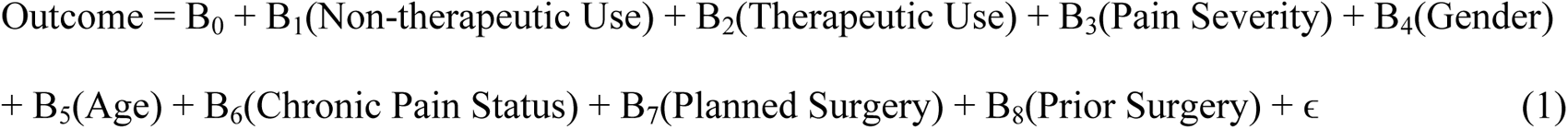

Past-week pain severity was treated as a continuous variable ranging from 0-10. Age was treated as a continuous variable, assessed per decade, starting at 19-29 years (1) up to 80+ years (7). Gender was treated as a categorical variable (Reference: Man). Pain chronicity was dichotomized into acute and chronic status (Reference: Acute, 6 months or more). Surgical treatment was represented by two indicator variables: planned surgery and prior surgery, with ’No surgical treatment’ serving as the reference group for both. Prior psychedelic use was represented by two indicator variables for non-therapeutic and therapeutic intent; participants with no lifetime history of psychedelic use served as the reference population. Significance was defined as *p*<.05. Diagnostic testing confirmed that the models met the assumptions of homoscedasticity (Breusch-Pagan Test, *p*<.05) and no multicollinearity (variance inflation factor for any independent variable < 5); although residuals deviated from normality (Shapiro-Wilk Test, *p*>.05), OLS regression was retained as it is robust to this violation in large samples via the Central Limit Theorem [38]. For all three models, we report R^2^, which ranges from 0 to 1, representing the proportion of variance in the dependent variable explained by the independent variables in a regression model. Higher scores indicate a better fit.

## RESULTS

### Participant Demographics

300 patients consented to participate. Five participants did not complete the survey because they were called into their appointment or otherwise did not have time. Therefore, 295 participants with musculoskeletal pain completed the survey. The age group in highest proportion was 60–69 years (26.9%, 79/294), followed by 70–79 years (24.8%, 73/294). Slightly more individuals identified as female (54.2%, 160/295) compared to male (45.4%, 134/295). Most participants had a post-secondary education; 41.2% (121/294) reported college or trade school education and 36.8% (108/294) reported a university degree. Thirty-eight percent (105/294) reported an annual household income ≥$100,000. Most of the participants identified as European/White (81.1%, 236/274). The patient demographics are described in **Table 1**.

**Table 1.** Participant Demographics.

| Variable | No. of patients (%) |
| --- | --- |
| Total N | 295 |
| **Age (N=294)** |  |
| 19–29 | 18 (6.1%) |
| 30–39 | 25 (8.5%) |
| 40–49 | 37 (12.6%) |
| 50–59 | 51 (17.3%) |
| 60–69 | 79 (26.9%) |
| 70–79 | 73 (24.8%) |
| 80+ | 11 (3.7%) |
| **Gender (N=295)** |  |
| Man | 134 (45.4%) |
| Woman | 160 (54.2%) |
| Non-binary | 1 (0.3%) |
| **Education (N=294)** |  |
| Less than high school | 6 (2.0%) |
| High school | 59 (20.1%) |
| College / trade school | 121 (41.2%) |
| Bachelor’s degree | 76 (25.9%) |
| Graduate degree | 32 (10.9%) |
| **Annual income (N=274)** |  |
| Less than \$25,000 | 23 (8.4%) |
| \$25,000–\$49,999 | 34 (12.4%) |
| \$50,000–\$74,999 | 56 (20.4%) |
| \$75,000–\$99,999 | 56 (20.4%) |
| \$100,000 or more | 105 (38.3%) |
| **Ethnicity (N=291)** |  |
| Black | 13 (4.5%) |
| East Asian or Southeast Asian | 8 (2.7%) |
| European/White | 236 (81.1%) |
| First Nations/Indigenous | 4 (1.4%) |
| Hispanic/Latinx | 3 (1.0%) |
| Middle Eastern | 10 (3.4%) |
| South Asian | 9 (3.1%) |
| Prefer to self-identify | 8 (2.7%) |

### Clinical Characteristics and Analgesic Use

Most participants reported chronic musculoskeletal symptoms, with 70.2% (198/282) experiencing pain for over six months (**Table 2**). Injury was the most common cause for MSK pain (51.3%, 143/279) and arthritis was second (40.5%, 113/279). Half of the participants (140/280) had previously undergone surgery for their condition and 12.1% (34/180) planned to undergo surgery. 73% of the cohort experienced high pain levels, where 36% (100/281) of participants reported moderate pain and 37% (105/281) reported severe pain.

**Table 2.** Clinical Characteristics.

| Variable | No. of patients (%) |
| --- | --- |
| **Duration of symptoms (N=282)** |  |
| Less than 6 months (acute) | 84 (29.8%) |
| 6 months or more (chronic) | 198 (70.2%) |
| **Major cause of your bone and joint pain (N=279)** |  |
| Injury (broken bone, dislocation, sprain) | 143 (51.3%) |
| Arthritis (joint pain) | 113 (40.5%) |
| Not sure | 15 (5.4%) |
| Other | 8 (2.9%) |
| **Underwent surgery for bone/joint condition (N=280)** |  |
| No | 106 (37.9%) |
| Not yet but I plan to | 34 (12.1%) |
| Yes | 140 (50.0%) |
| **Pain severity (0 = no pain, 10 = extreme pain) (N=281)** |  |
| Mild (0–3) | 76 (27.0%) |
| Moderate (4–6) | 100 (35.6%) |
| Severe (7–10) | 105 (37.4%) |

In the sample, analgesic use was common, with 74.6% (209/280) of participants reporting use of medications for pain (**Table 3**). Over the counter medications were the most frequently used, (48.1%, 142/208), followed by opioids (29.2%, 86/208) and non-opioid prescription medications (22.0%, 65/208). Cannabis use was reported by 9.8% (29/208) of participants while 4.3% (9/208) reported using psychedelics for pain. The perceived effectiveness of analgesics varied. Opioids were rated highest (mean 5.86/10), followed by cannabis (5.45/10) and then over the counter agents (5.00/10) and psychedelics (4.67/10).

**Table 3.**
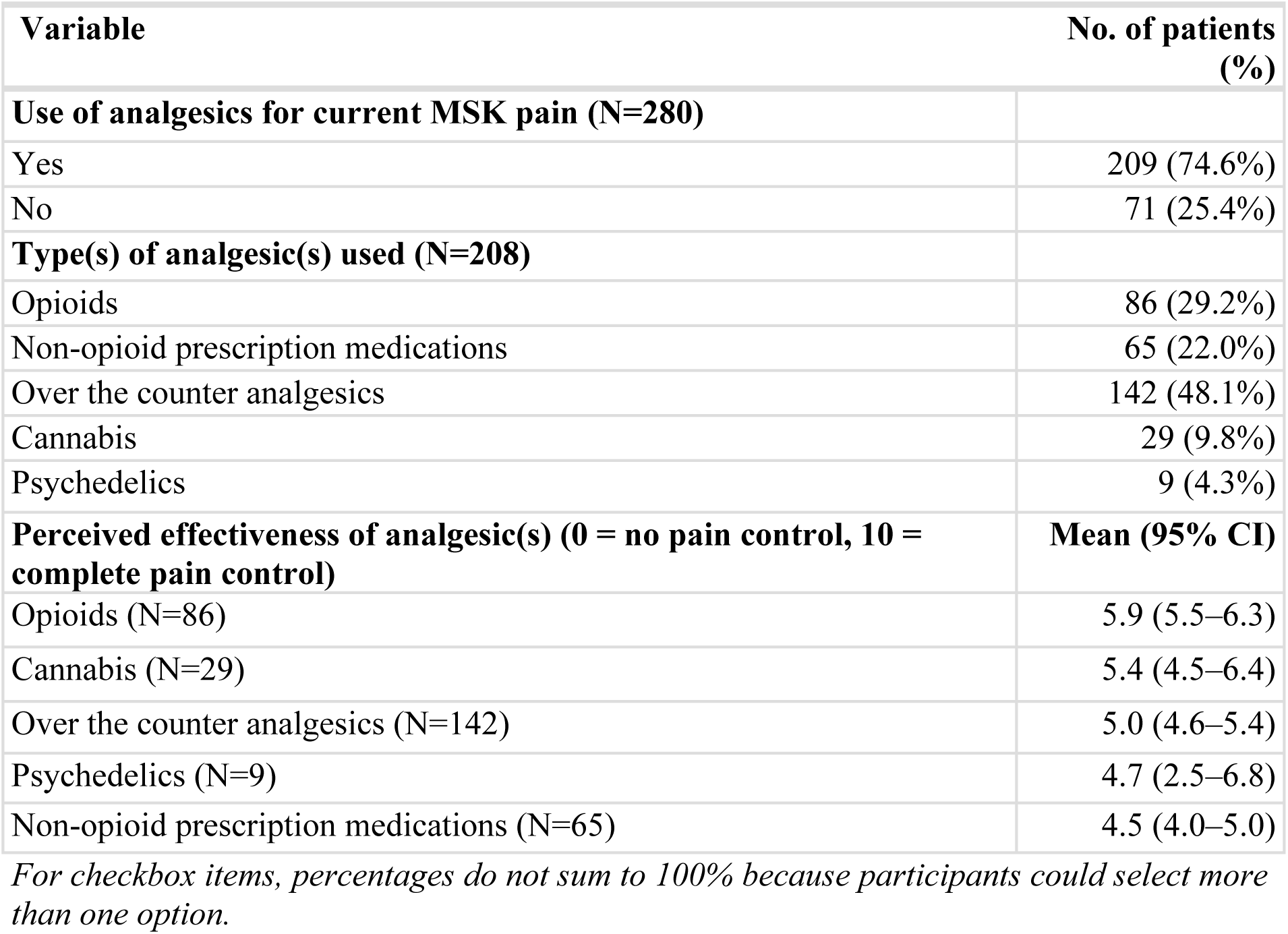
Analgesic Use.

### Psychedelic use

Almost one fourth of participants (23.5%) reported prior psychedelic use (**Table 4**). Most use was non-therapeutic (17.7%, 52/294), and only 2.0% (6/294) of participants reported therapeutic reasons for use exclusively, while 3.7% (11/294) of participants reported both. Among the individuals that reported therapeutic use of psychedelics (n = 17), mental health and chronic pain (both 47.1%, 8/17) were the most common indications. Non-therapeutic motives (84.4%, 54/64) were most reported among people using psychedelics recreationally, followed by personal growth (23.4%, 15/64) and spiritual/physiological motives (18.8%, 12/64). Thirty-two percent (22/69) of people using psychedelics reported improved health and well-being, 10.1% (7/69) reported worsened health, and the majority reported no change (58.0%, 40/69).

**Table 4.** Psychedelic Use.

| Variable | No. of patients (%) |
| --- | --- |
| **Prior psychedelic use (N=294)** |  |
| None | 225 (76.5%) |
| For therapeutic reasons only | 6 (2.0%) |
| For non-therapeutic reasons only | 52 (17.7%) |
| For both therapeutic and non-therapeutic reasons | 11 (3.7%) |
| Past-year therapeutic use | 8 (2.7%) |
| **Therapeutic reasons for psychedelic use (N=17)** |  |
| Acute pain | 2 (0.7%) |
| Chronic pain | 8 (2.7%) |
| Headaches | 1 (0.3%) |
| Mental health | 8 (2.7%) |
| Other | 3 (1.0%) |
| **Non-therapeutic reasons for psychedelic use (N=64)** |  |
| Spiritual/psychological | 12 (4.1%) |
| Personal growth/personal exploration | 15 (5.1%) |
| To manage past trauma | 7 (2.4%) |
| For fun | 54 (18.3%) |
| Social peer pressure | 9 (3.1%) |
| Boredom/just to try it | 14 (4.7%) |
| Other | 2 (0.7%) |
| **Psychedelics' overall effect on health and wellbeing (N=69)** |  |
| Made health and well-being better | 22 (31.9%) |
| No change in health and well-being | 40 (58.0%) |
| Made health and well-being worse | 7 (10.1%) |

*For checkbox items, percentages do not sum to 100% because participants could select more than one option. Additional therapeutic reasons reported via free text included Cesarean section, back pain, and "party therapy" (reported verbatim). Additional non-therapeutic reasons included "teenager" (reported verbatim), with one participant declining to respond.*

Psilocybin was the most commonly used psychedelic for both therapeutic (69.2%, 9/13) and non-therapeutic (79.0%, 49/62) reasons (**Fig 1**). Ketamine (23.1%, 3/13) was the second most common therapeutically used psychedelic. LSD was the second most common non-therapeutic substance (51.6%, 32/62) followed by MDMA (29.0%, 18/62) and ketamine (11.3%, 7/62). Four participants who reported therapeutic use and one participant who reported non-therapeutic use did not select any substances from the provided checklists.

**Fig 1.**
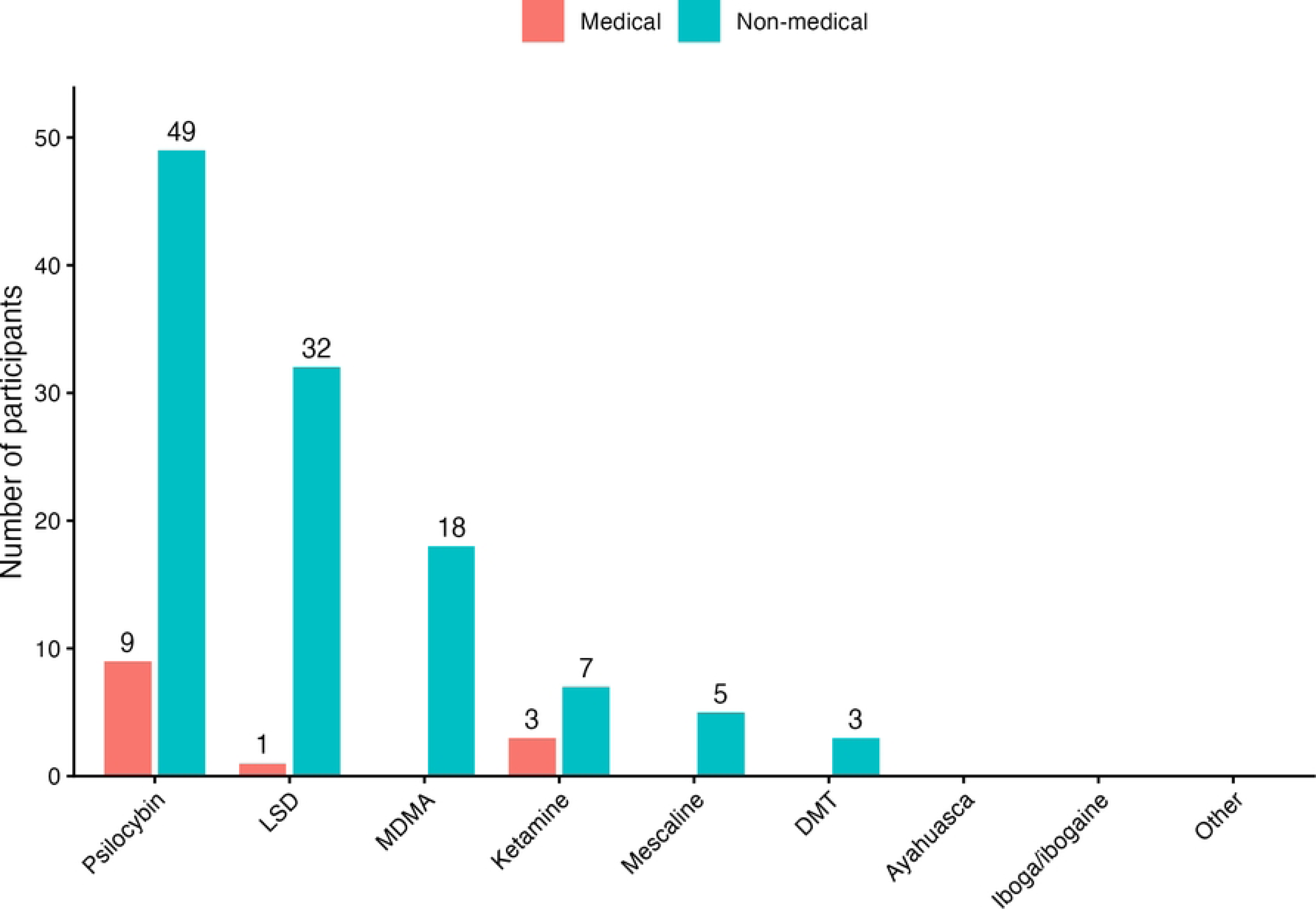
**Psychedelic Substances Used for Therapeutic and Non-therapeutic Reasons. Note: Participants could select more than one option.**

### Perceived Potential, Willingness, Comfort, and Perceived Risk

As seen in **Fig 2**, nearly half of the participants (45.5%, 132/290) reported high comfort discussing psychedelics with a healthcare professional; however, only 10 participants had done so. Among these participants, discussions were generally positive (mean 3.70/5). The perceived analgesic potential of psychedelics was most frequently rated as moderate (49.3%, 137/278) or high (30.6%, 85/278).

**Fig 2.**
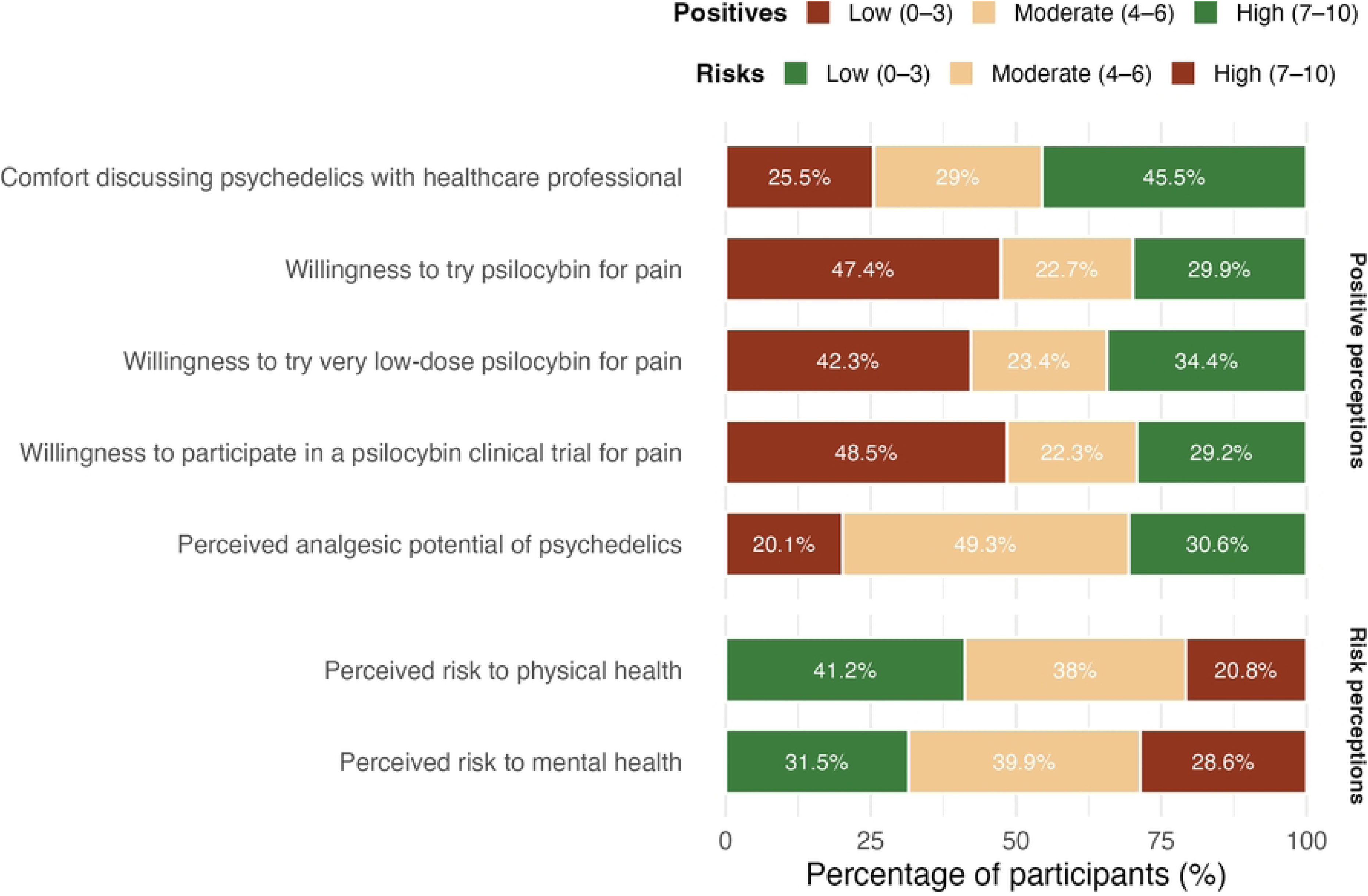
**Attitudes and Risk Perceptions.**

However, almost half of respondents indicated low willingness to try psilocybin as a treatment (47.4%, 138/291) or to participate in a clinical trial (48.5%, 141/291). Willingness improved when considering microdose psilocybin, with 34.4% (100/291) reporting high willingness. Perceived risk to physical health was predominantly rated as low (41.2%, 115/296), while perceived risk to mental health was most frequently rated as moderate (39.9%, 110/276).

Overall, participants expressed a small but significantly higher willingness to try microdose versus macrodose psilocybin for pain management (mean difference = 0.47 points on a 10-point scale; *p*<.001). The perceived risk of psychedelics (in general) to mental health was significantly higher than physical health (mean difference = 0.65, p<.001; **S1 Table**). As seen in **Fig 3**, individuals with prior psychedelic experience demonstrated more favorable attitudes toward the therapeutic use of psychedelics compared to those who did not report prior use. Individuals who had used psychedelics reported greater comfort discussing psychedelics, a stronger belief in their analgesic potential, and a higher willingness to both try psilocybin and participate in clinical trials (all *p*<.001). Furthermore, this group perceived psychedelics to have fewer physical and mental health risks (both *p*<.001; **S2 Table**).

**Fig 3.**
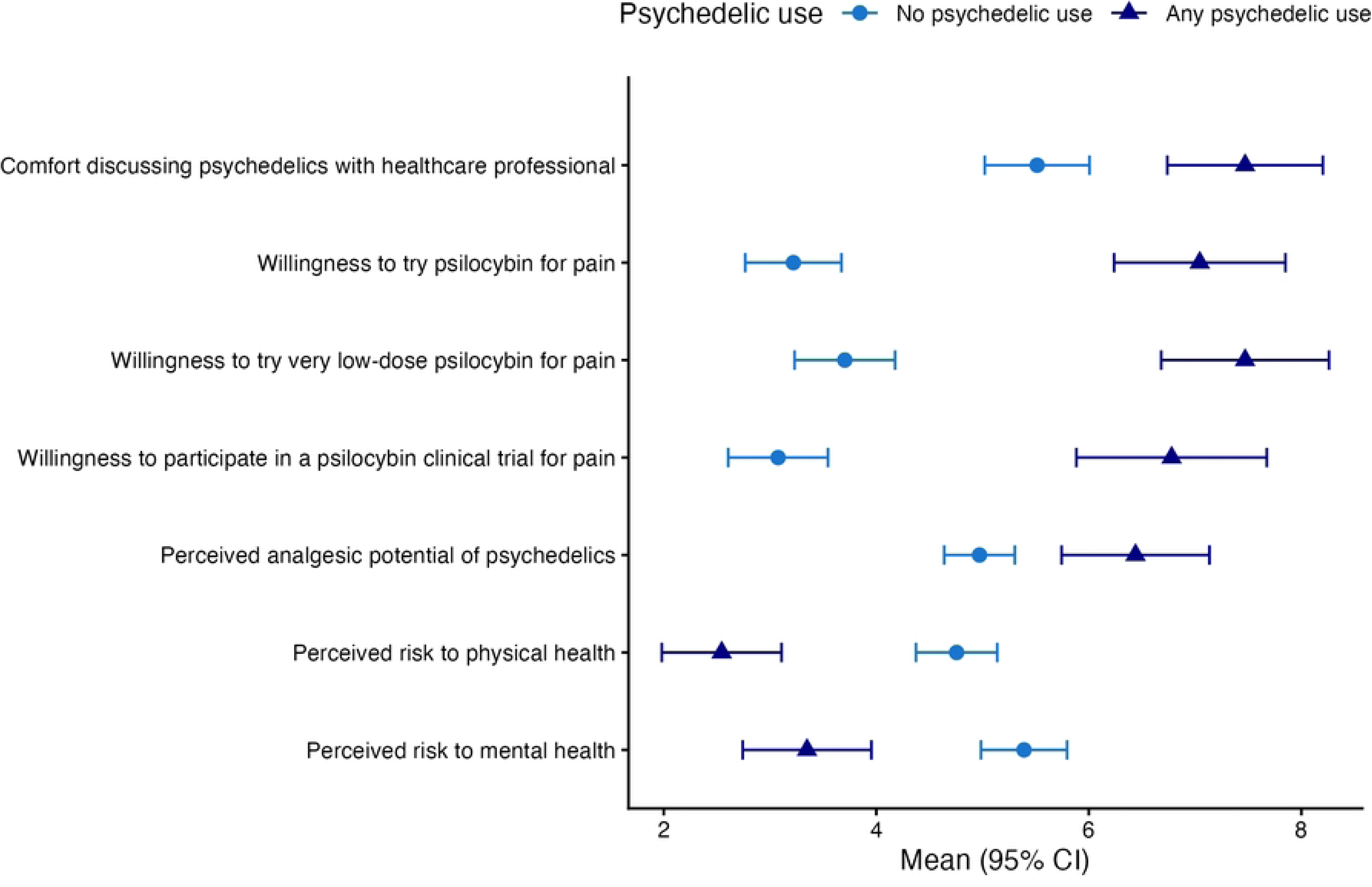
Attitudes and Risk Perceptions Stratified by Psychedelic Use.

As seen in **Fig 4**, individuals with moderate-to-high pain ratings compared to low pain ratings were significantly more willing to try psilocybin for pain, both at a macrodose (mean difference=1.23, *p*=.0166) and microdose (mean difference=1.21, *p*=.0145), and more willing to participate in a clinical trial (mean difference=1.46, *p*=.0083), but exhibited no significant differences in comfort levels, perceived analgesic potential, or risk perceptions **(S3 Table)**.

**Fig 4.**
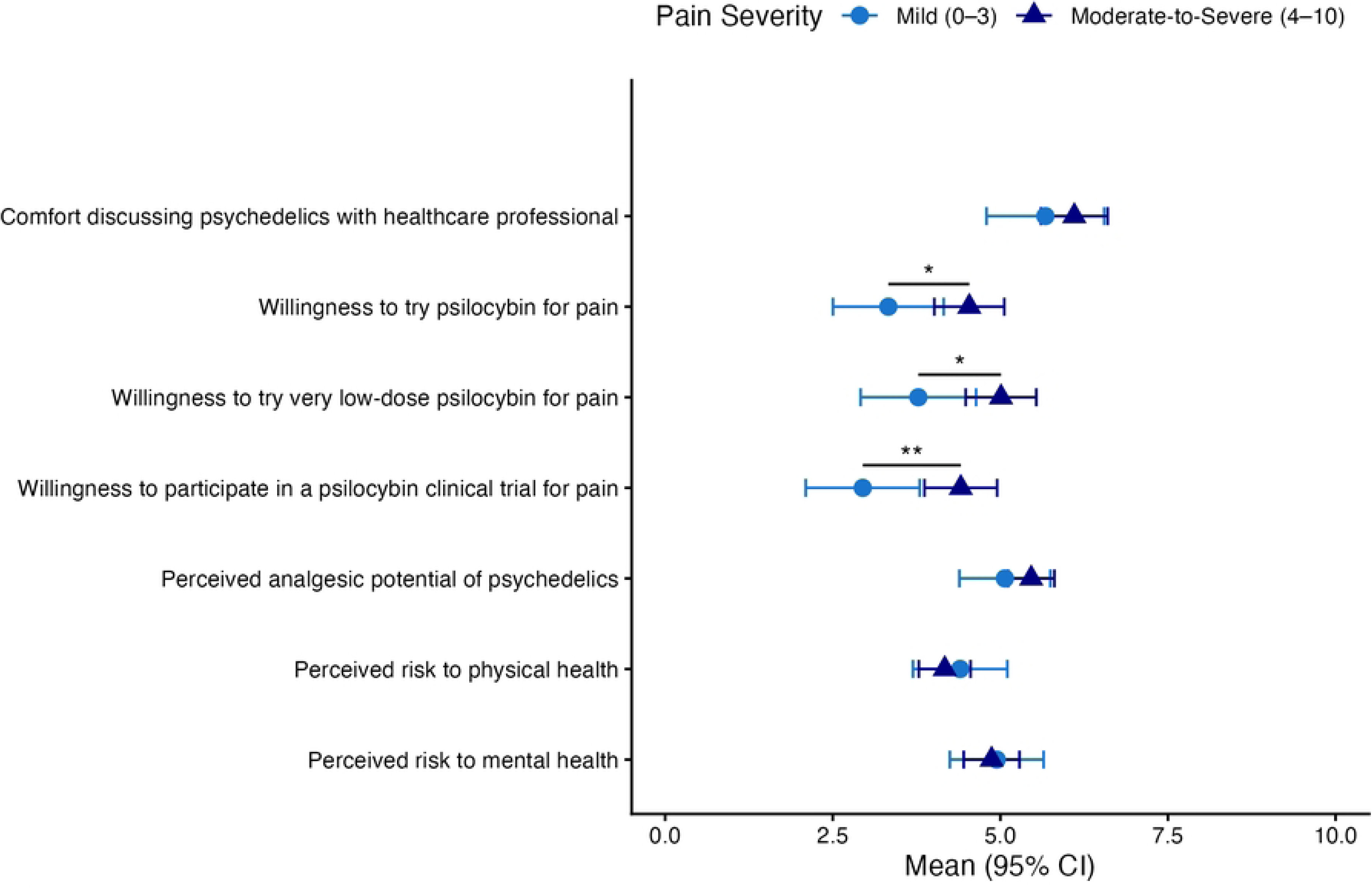
Attitudes and Risk Perceptions Stratified by Pain Severity.

As shown in **Table 5**, linear models revealed that the best-fitting model, examining willingness to try a macrodose, only explained up to 23% of the variance in attitudes. Models predicting willingness to try psilocybin were more robust (e.g., Willingness to try; *R^2^*=0.23, *F*(8, 259)=9.88, *p*<.001, adj. *R^2^*=0.21; microdose willingness; *R^2^*= 0.21, *F*(8, 259)=8.77, *p*<.001, adj. *R^2^*=0.19) than the model predicting analgesic potential (*R^2^*=0.07, *F*(8,259)=2.41, *p*=0.016, adj. *R^2^*=0.04).

**Table 5.** Multivariable Regression Models for Patients’ Perceptions of Therapeutic Psychedelics.

| Variable | Analgesic Potential |  | Willingness to Try |  | Willingness to Try Microdose |  |
| --- | --- | --- | --- | --- | --- | --- |
|  | *B* (SE)<br>[95% CI] | *p* | *B* (SE)<br>[95% CI] | *p* | *B* (SE)<br>[95% CI] | *p* |
| Prior non-therapeutic psychedelic use | 1.05 (0.42)<br>[0.22, 1.87] | **.013\*** | 3.16 (0.55)<br>[2.08, 4.24] | **<0.001\*** | 2.82 (0.56)<br>[1.71, 3.93] | **<0.001\*** |
| Prior therapeutic psychedelic use | 1.18 (0.76)<br>[-0.31, 2.67] | .119 | 2.42 (0.99)<br>[0.48, 4.37] | **.015\*** | 2.48 (1.02)<br>[0.48, 4.48] | **.015\*** |
| Prior surgery (vs none) | -0.49 (0.35)<br>[-1.17, 0.2] | .162 | -1.12 (0.45)<br>[-2.01, -0.23] | **.014\*** | -1.06 (0.47)<br>[-1.98, -0.14] | **.024\*** |
| Past-week pain severity | 0.05 (0.07)<br>[-0.08, 0.18] | .436 | 0.21 (0.09)<br>[0.04, 0.37] | **.017\*** | 0.2 (0.09)<br>[0.02, 0.37] | **.027\*** |
| Age category (per decade) | -0.09 (0.11)<br>[-0.3, 0.13] | .420 | -0.21 (0.14)<br>[-0.48, 0.07] | .147 | -0.3 (0.14)<br>[-0.58, -0.01] | **.042\*** |
| Female (vs male) | -0.32 (0.33)<br>[-0.97, 0.33] | .328 | -0.66 (0.43)<br>[-1.51, 0.19] | .125 | -0.73 (0.44)<br>[-1.6, 0.14] | .102 |
| Chronic pain > 6 months (vs acute) | -0.06 (0.38)<br>[-0.81, 0.69] | .880 | 0.28 (0.5)<br>[-0.7, 1.25] | .580 | 0.33 (0.51)<br>[-0.67, 1.34] | .515 |
| Planned surgery (vs none) | -0.65 (0.54)<br>[-1.71, 0.41] | .230 | -0.87 (0.7)<br>[-2.26, 0.52] | .218 | -1.28 (0.72)<br>[-2.71, 0.14] | .078 |

Non- therapeutic psychedelic use was a robust predictor across all models. Specifically, prior non-therapeutic use was associated with an increase of 1.05 points on the 10-point analgesic potential scale (*p*=.013) and increases of 3.16 and 2.82 points on the 10-point willingness scales for macrodoses and microdoses, respectively (*p*<.001). Prior therapeutic psychedelic use showed similarly substantive associations with willingness to try both macrodoses (*B*=2.42, *p*=.015) and microdoses (*B*=2.48, *p*=.015). Prior surgical treatment was a negative predictor of willingness to try macrodoses (*B*=-1.12, *p*=.014) and microdoses (*B*=-1.06, *p*=.024). In contrast, while increased pain severity (per 1-point increase on a 0–10 scale) and younger age (per decade) were statistically significant, their coefficients (*B*=0.20 to 0.21 for pain; *B*=-0.30 for age) suggest these factors have lower relevance relative to the impact of prior experience with psychedelics.

Concern over health risks was the most reported barrier to participating in a clinical trial (32.5%,). Adequate pain management with current therapy (23.4%), financial/employment/transportation constraints (16.9%) and the legal status of psychedelics (5.6%) were additional barriers to trial participation. 26.4% of participants said that none of the obstacles on the list would keep them from participating **(Fig 5)**.

**Fig 5.**
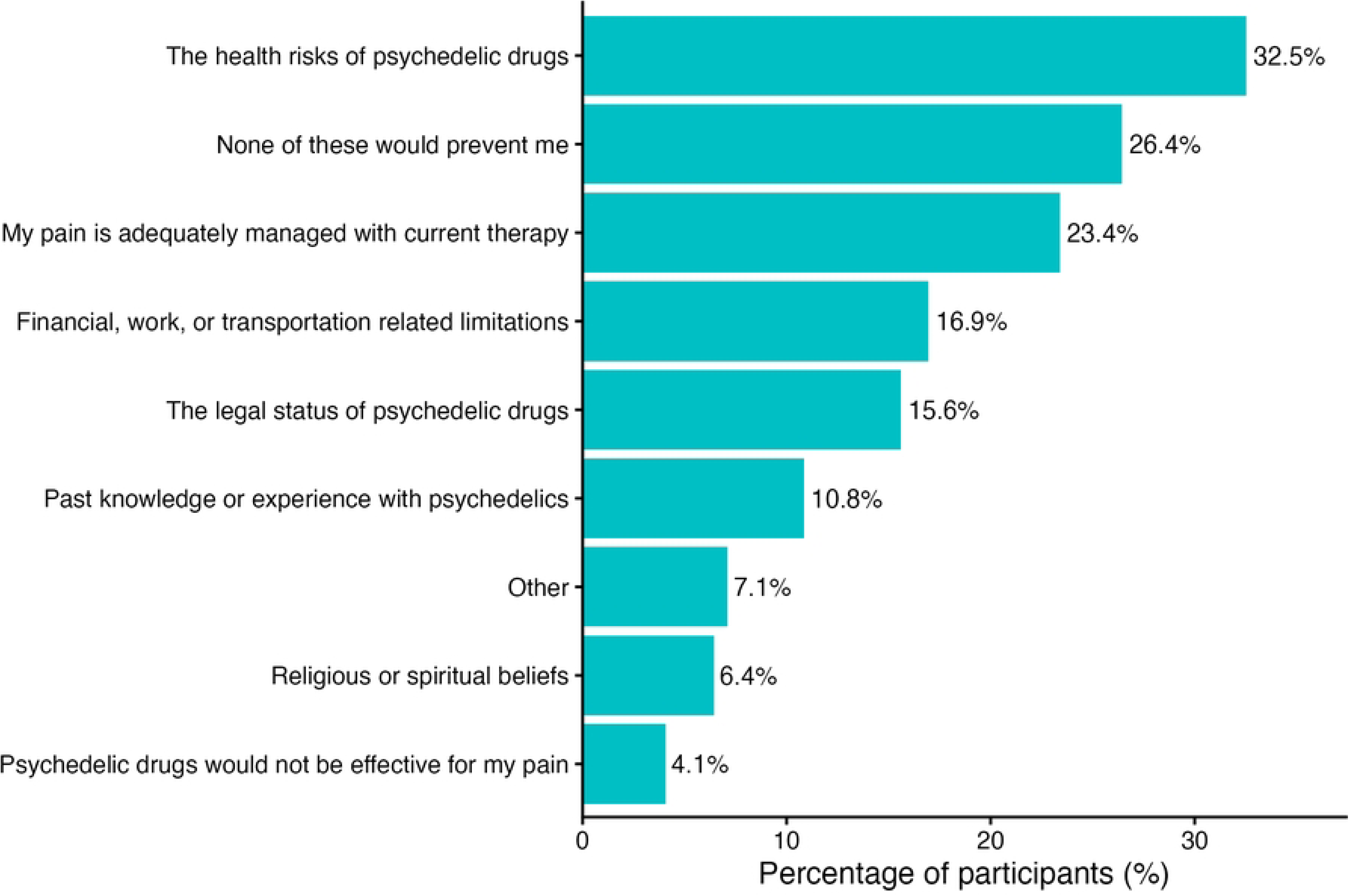
Barriers to Participating in a Psilocybin Clinical Trial. Note: Participants could select more than one option.

## DISCUSSION

Our survey of patients attending an orthopaedic surgery clinic found almost one in four had prior experience with psychedelics. Further, the majority of patients believed that psychedelics were effective for pain relief and were somewhat willing to participate in clinical trials evaluating psychedelics, particularly when administered as a microdose.

Our finding that nearly a quarter of our respondents reported prior psychedelic use is consistent with Glynos et al. where 30% of participants with fibromyalgia reported past use [25]. This comparison is highly relevant given that musculoskeletal pain is a defining symptom of fibromyalgia. The demographic divergence between these two samples makes these similar prevalence rates compelling. While the sample in the Glynos et al. survey was overwhelmingly female (91.5%), aligning with epidemiological trends in fibromyalgia, our orthopaedic clinic sample was gender-balanced (54.2% female). Perceived changes in health and well-being because of psychedelics were mostly neutral (58.0%), with 31.9% reporting positive impacts of psychedelic use and 10.1% reporting negative impacts. This aligns with population-based studies, which have found mixed, but mostly positive or null associations between psychedelic use and mental [39–42] or physical health [43]. However, observational data on naturalistic psychedelic use has found self-perceived improvements in overall well-being [44], positive health behavior changes [45,46], and better chronic pain [47] functioning across both general and clinical populations. Despite the high prevalence of use and neutral or positive outcomes, only 10 participants had discussed psychedelics with a healthcare professional.

A primary finding of this study is the nuanced willingness of MSK patients to engage with psychedelic treatment. While nearly half of the cohort exhibited baseline reluctance toward macrodose psilocybin or clinical trial participation, acceptance increased significantly when considering a microdose option. Willingness was shaped by clinical history: greater pain severity predicted greater willingness, while prior surgical intervention was as a negative predictor. This indicates patients in a refractory state, with high symptom burden and who have not undergone a definitive surgical solution, may be most motivated to try this experimental treatment. This finding presents an ethical challenge for future trial design: the individuals who are most motivated to participate are also those who likely face the greatest physical and logistical barriers to doing so. Given the current paucity of clinical evidence for psychedelics in musculoskeletal pain, protocol designs must carefully account for patient vulnerability. Severe pain conditions can severely limit mobility and the ability to tolerate the prolonged, sedentary in-person sessions required by standard psychedelic dosing protocols. Beyond clinical status, younger age predicted willingness for low-dose treatment, perhaps reflecting a greater familiarity with the recently emerging practice of microdosing [48,49]. Ultimately, and consistent with studies on attitudes towards psychedelics in clinical populations, personal experience was the strongest predictor of positive perceptions of psychedelics [50].

Regarding barriers to clinical trial participation, concern over health risks was the most frequently cited. This apprehension is predominantly psychological; participants reported significantly higher perceived risks to their mental health than to their physical health. This may explain respondents’ preference for microdoses of psilocybin, which aim to provide therapeutic benefit while circumventing intense psychoactive effects. A quarter of our participants identified adequate pain management with current therapy, reflecting disinterest in pursuing alternative treatment. Finally, logistical constraints, including financial, employment, and transportation hurdles, was the second most common barrier to use of psychedelics for pain management. Addressing these issues, for example through decentralized trial designs (e.g., through take-home doses) or participant stipends, will be essential for ensuring the feasibility of future trials.

### Strengths and Limitations

This study has several notable strengths. To our knowledge, this anonymous cross-sectional survey of 295 orthopaedic clinic patients is the first to describe prior psychedelic use, attitudes, and willingness to try psilocybin for pain management among individuals with MSK disorders. Multivariable modeling allowed us to control for specific clinical variables to identify independent predictors of willingness.

Limitations include a cross-sectional design that precludes causal inferences, and a reliance on self-reporting, which risks recall and social desirability biases regarding scheduled substances, although the survey was anonymous to reduce potential bias. Additionally, the predominantly older, white, single-center cohort may limit generalizability to broader populations.

### Significance and Future Recommendations

Patients with severe MSK pain are open to alternative therapies like psychedelics, yet significant safety concerns remain. We recommend that healthcare providers lead proactive, non-judgmental conversations when patients indicate interest in psychedelics for pain and inform patients about current and future clinical trials. During trial recruitment, concerns related to the legality and safety of psychedelics can be addressed in the consent process by emphasizing safety measures, medical oversight, and legal protections [51]. The process should explain that investigational products are manufactured in compliant facilities and administered in controlled environments approved by Health Canada.

The logistical constraints cited by our patients are particularly relevant to full-dose psychedelic assisted therapy trials which demand exceptionally high time commitments, including full-day dosing sessions, that may prove physically and logistically prohibitive. Recently completed and ongoing clinical trials investigating psilocybin for pain conditions employ diverse dosing strategies, ranging from high-dose psilocybin-assisted therapy [28,52–57], to lower-to-moderate non-psychedelic doses [22,23,58–60], to comparative designs comparing high and low doses [29]. While future research must ultimately evaluate both microdose and macrodose protocols to determine if the subjective psychoactive experience is a necessary mediator of therapeutic efficacy, a microdosing framework appears to be perceived as a more appealing target for initial investigation of the potential of psilocybin for MSK pain.

## CONCLUSIONS

Study findings suggest most patients hold neutral to positive perceptions of psychedelics for pain, with over three quarters of the participants reporting moderate or high perceived analgesic potential of psilocybin and over half reporting moderate to high willingness to participate in a psilocybin clinical trial. Addressing perceived barriers, including risk of harms, practical barriers to trial participation, and gaps in patient knowledge, should be considered when designing future trials.

## Data Availability

This dataset contains sensitive data involving potentially illicit activities, therefore the dataset is available from the corresponding author on reasonable request upon obtaining appropriate institutional ethics approval and completing a data sharing agreement.

## ACKNOWLEDGEMENTS

We thank the patients at the orthopaedic clinic of St. Joseph’s Healthcare Hamilton for volunteering in our study.

## SUPPORTING INFORMATION

**S1 Appendix. Survey items.**

**S1 Table. Within-subjects comparison of attitudes.**

**S2 Table. Independent comparisons of attitudes stratified by prior psychedelic use.**

**S3 Table. Independent comparisons of attitudes stratified by low and moderate-to-severe pain.**

